# Pitch Reaction Test - Stepping towards a novel measure of batting cognition

**DOI:** 10.1101/2025.06.05.25329069

**Authors:** Tiago Lopes, Samantha Kerr, Nabeel Vandayar, Benita Olivier

## Abstract

Little is known about the cognitive acuity of batters in cricket. Potentially this data is limited due to the lack of practical methods of assessing cognitive function congruently while batting. In this study we describe the development of a choice reaction task paradigm which is deployed on a cricket pitch. The Pitch Reaction Test (PRT) analogues the initial central processing and motor response of stroke selection to quantify psychomotor performance. A prototype PRT was assessed in a laboratory to model its measurement accuracy against reviewed slow-motion footage of motor responses from trials of 23 participants. Linear models compared PRT performance vs an established computer-based task and user anthropometry. Measurements obtained from the PRT were strongly correlated with review of video footage (***r* = 0.97 − 0.99**), although Bland-Altman analysis reveals a linear trend of differences increasing with increases in mean response times. Neither participant height (***β* = −0.01, *p* = 0.54**) or leg-length (***β* = 0.0, *p* = 0.58**) were predictors of PRT performance. Choice response performance on an equivalent computer assessment has a weak (***R^2^* = 0.16**) negative relationship (***β* = −0.21, *p* = 0.04**). Overall, the PRT demonstrates a novel solution to measuring sport specific psychomotor function in cricket enabling multiple avenues of future research.

## 1 Introduction

The cognition of cricket batters has not been extensively studied and authors have questioned if data from standardised, cognitive component ?pen and paper? assessments can be generalised to describe sport-specific performance [1]. Another approach attempts to quantify a sport-specific cognitive parameter by mimicking the stimuli, temporal constraints, and information processing stream of match-play. For example, Furley and colleagues [2] assessed visual awareness by presenting game scenarios depicting players are available or unavailable to receive a pass. The task paradigm enabled the authors to manipulate and discuss how the contents of the athletes working memory may bias the athletes? attentional mechanisms to seek similar contents in the current visual field. Importantly, such data provides objective practical implications to the performance enhancement and coaching environment.

In cricket, a parameter of cognition critical to batting performance is psychomotor function [3, 4]. That is, the cognitive processing of visual and perceptual stimuli coupled with a complex movement behaviour [5]. The extreme temporal constraints of intercepting a high-velocity cricket ball requires substantial processing speed of specific contextual stimuli (bowling action, ball flight). Coupled with precise, rushed movement responses ? Sport Specific Psychomotor Function (SSPF) [6]. Therefore, a method of quantifying SSPF, mimicking sensory-motor characteristics of batting, may provide much needed data for research with practical insights for the game.

The aim of the current study is to propose a task paradigm and design to quantify SSPF in cricket. The chosen task paradigm is analogous to established assessments of psychomotor function ? response time (milliseconds) to distinguishable stimuli with corresponding distinct inputs/responses [5]. The design provides a means of conducting assessments on a cricket pitch, concurrently with batting. The distinguishable stimuli, in the form of coloured lights, mimic the potential locations that deliveries will impact and bounce. The corresponding movement response is to initiate a ?front foot? Or ?back foot? stroke by completing either a forward or backward lunge. A prototype device is developed and tested for recording accuracy against a high-speed camera.

## 2 Methods and Materials

### 2.1 Pitch Reaction Test - Background

In one trial of the PRT, the subject assumes the posture of a batter ? knees slightly bent and shoulders parallel to the light stimuli placed lengthwise down the artificial pitch. In this starting position, the subject places both feet in a defined position demarcated on the turf. The ‘front foot’ is placed closest and perpendicular to the light stimuli and the ‘back foot’ placed similar but distal from the light stimuli by approximately shoulder width. The light stimuli mimic the incoming direction of deliveries from the bowler and two distinct categories or locations where the delivery will bounce. In cricket, a delivery can be coarsely categorised based on the distance a delivery impacts the pitch causing it to bounce in front of the batter. The height of a given delivery when it reaches the batter is therefore determined by the location of bounce. Deliveries which bounce further from the batter (called ?short? deliveries) have more length of the pitch to travel and gain incline compared to deliveries which bounce close to the batter (?full?) and are therefore intercepted before regaining much height. Importantly, the category of delivery, either short or full, determines the primary movement the batter must complete to successfully intercept the delivery. Short deliveries tend to reach the batter at above chest height and ideally should be redirected back towards the ground requiring the batter to remain fully upright at interception. In contrast, for a full delivery, the batter will move towards the location of bounce, ideally intercepting the delivery low to the ground, smothering the potentially unpredictable bounce.

Using this concept, the PRT displays a stimulus at either a full length or short length then assesses the latency of the batter/subject?s decision and movement response to the stimuli. A simple, dependable circuit was designed to 1) display a light stimulus (green light) at one of two locations along the length of a cricket pitch and 2) measure the latency in milliseconds between stimulus presentation and user-input/response at the users? feet. Software on and delivered to a microprocessor (Arduino, Ivrea, Italy) oversees the randomisation of circuit behaviour i.e., which location displays a green light and the delay between successive trials/stimuli. An outline schematic of the device and circuitry is drawn in 1.

### 2.2 User Input or Instructions

For short deliveries, the light stimuli most distal from the subject is activated ? to respond the subject?s backfoot must move from its starting position further backward to a defined location demarcated on the turf keeping the front foot in contact with the ground at its starting position. For full deliveries, the light stimulus closest to the batter is activated, to respond the front foot moves forward to a defined location, backfoot remaining in contact with the ground.

### 2.3 Hardware

The PRT can be sub-divided into three functional units. The first unit being an array of four 650nm 5mw red dot lasers (Electronics International Enterprise, distributed by ManTech Electronics, South Africa) connected in series to an independent source of 5V direct current. The lasers are mounted onto circuit boards and fixed to a wooden rail at predefined distances corresponding to the starting and response-ending positions of the front/backfoot (Figure 1). The second unit is a complementary sensory array of four, 20mm diameter, photosensitive variable resistors (Senba Optoelectronic, Shenzen, China). Mounted to circuit boards and fixed to a second wooden rail, the sensor height from the ground is 0.5cm. The variable resistors are semiconductive materials with a peak response sensitivity to 500-700nm light. The sensory and laser array are placed parallel 2m apart and each laser points perpendicularly to the opposite wooden rail towards its paired variable resistor on the sensory array. The third functional unit is connected to the sensory array via Network LAN cat6 cable. One LAN cable from each photosensitive resistor connects to four analogue pins of the Arduino Nano (Arduino, Ivrea, Italy) mounted on a circuit board. The output of the Arduino Nano is also delivered via LAN cat6 cable, instead using three digital pins to control one LED. Two LEDs were controlled by the Arduino Nano, one placed 2.5m from the starting ‘front foot’ position and the second 6m from this position. Therefore, the third functional unit can be conceptualized as the processing and control unit -processing signal from the complimentary laser-sensory arrays and controlling the behaviour/activity of the two LEDs.

**Fig 1.**
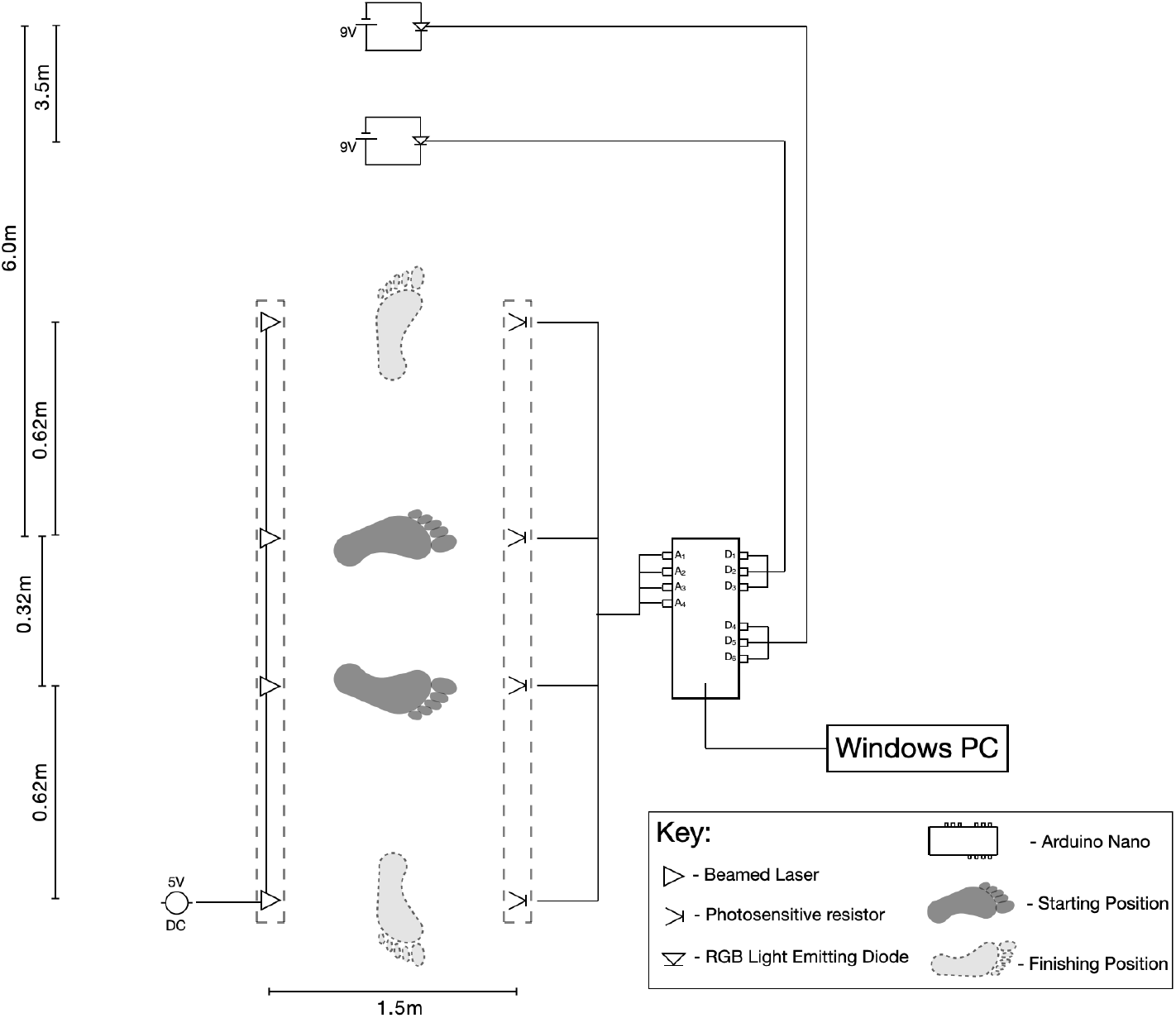
Schematic of Pitch Reaction Test componentry and layout - not drawn to scale

**Fig 2.**
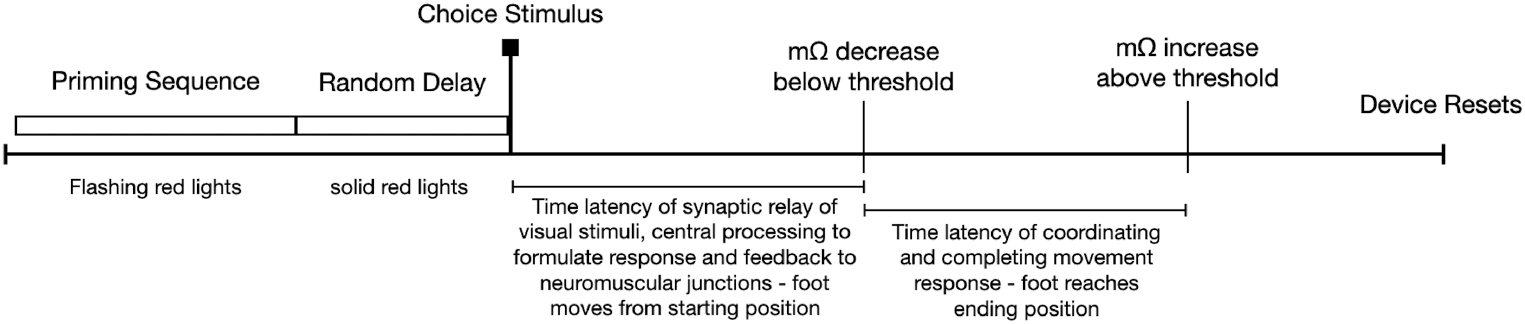
Sequence of events and measurement cut-offs at starting and ending sensory arrays

The PRT analyses movement of the users’ feet by measuring total resistance (mΩ) at the four circuits of the sensory array. When exposed to direct, bright light the total resistance applied by the photosensitive resistor to its circuit is reduced by 600-700 mΩ. When a user stands at the front and backfoot starting positions, the corresponding photosensitive resistors are occluded from their paired lasers by the users? feet/lower limbs. The remaining two sensory circuits receive unimpaired light from their corresponding lasers and therefore total resistance measured at these circuits is 600-700 mΩ less than at the occluded circuits of the starting position. The photosensitive resistors are validated to respond to changes in lux (light brightness) in 30ms. When responding to a stimulus, the user’s foot will move from the starting position, providing the corresponding sensory circuit with unimpaired light, and finish at the demarcated end position on the turf ? occluding its corresponding sensory circuit, increasing total resistance by 600-700 mΩ.

### 2.4 Software

The functioning of the PRT is enabled by two scripts written in Arduino Programming Software (Arduino, Ivrea, Italy) and Python programming language v3.1 [7]. To summarize, the Python script entails the following instructions/functions; i) generate a random integer between 0 and 1 from a binomial distribution, ii) convert the integer to a character string of ‘front’ (> 0 x ≤ 0.5) or ‘back’ (> 0.5 x ≤ 1.0), iii) return one random letter between A-G from a polynomial distribution, iv) send combination of generated character string and letter (e.g. frontA) to Arduino, v) compile signal returned from the Arduino into a .txt file.

The output from the Python code is delivered to the Arduino Nano and the microprocessor is initialized with a script containing the following instructions/functions; i) intermittently power both LEDs to display red light in a sequence of 100ms powered and 1500ms rest ? the rest interval is then halved every cycle providing the impression of increasing speed/frequency, this is the priming sequence, ii) power both LEDs to display red light for a defined period by interpreting the received signal where A = 400ms, increasing in 200ms increments till G = 1600ms, iii) interpret character string from received signal to power corresponding LED circuit with green light and rest the alternate LED circuit iv) calculate the interval between powering the LED circuit and total resistance reaching threshold at the corresponding sensory circuit (front or back foot starting position), v) calculate the interval of reaching resistance threshold between the starting and finishing position. vi) send both calculated intervals to the connected windows PC.

Thresholds of total resistance were defined to consistently label events occurring at the sensory circuits where photosensitive resistors have a response time of 30ms and total resistance is continuously measured at 1000*Hz*. Therefore, at sensory circuits of the starting positions where sensors are occluded the threshold was labelled as < 200*m*Ω and > 600*m*Ω for the sensory circuits at ending positions. Hence, two variables are obtained from one trial of the PRT. One measurement of the interval between stimulus presentation and primary responsive movements - this can be regarded as pre-motor time i.e., time to register, process and respond to stimuli. And a second measurement of the interval between first movement and completion of movement/response. The summation of both measures can be regarded as choice-response motor time.

A calibration function in the Arduino code provides a live trace of output from the sensory circuits. The live trace (Arduino IDE serial plotter) was used to verify that each sensory circuit reached threshold in obscured and unobscured conditions ensuring no interference is caused by ambient light. The calibration script was run before every subject was assessed.

### 2.5 Procedural Overview

The measuring accuracy of the developed device was assessed in a single-assessment laboratory study. The study was conducted in The Movement Physiology Research Laboratory at the University of the Witwatersrand, South Africa. The Pitch Reaction Test (PRT) was set-up on an artificial turf with length = 12 meters and width = 3m under artificial lighting. In the quiet laboratory, a PC was set-up for participants to perform a seated cognitive assessment of response times with a task domain akin to the PRT. Healthy males (n = 10) and females (13) were recruited within an age range of 18-30 years. In approximately 1 hour, participants were habituated to the PRT, performed the computer-based cognitive assessment, and thereafter the final assessment using the PRT. Anthropometry was also collected prior to these assessments.

#### Ethics Statement

The aims and procedures of the study were carefully explained to participants who thereafter signed voluntary informed consent. All procedures were reviewed by the Human Research Ethics Committee (clearance no: M171165)

To habituate participants to the PRT, the device was initiated to run continuously during the demonstration. Participants performed an undefined number of trials until satisfactory task proficiency, judged as 15 consecutive correct responses, was demonstrated. Assessing the learning response to the PRT was beyond the scope of this experiment, therefore satisfactory task proficiency was deemed on ability to provide data to validate the recording accuracy of the PRT and not a required threshold of performance. For the final PRT assessment, a camera (GoPro Hero8, San Mateo, USA) was set-up 8cm above the artificial turf capturing view of both light stimuli and lower limbs of the participant. Before starting the final PRT assessment, the camera was initiated to record at 240 frames per second for the entirety of the assessment.

### 2.6 Anthropometry

Stature was recorded to the nearest 0.5cm using a stadiometer. True leg length [8] was recorded to the nearest 0.5cm using a tape measure placed at the bony prominence of the iliac crest and medial malleolus. Both legs were measured to obtain an average.

### 2.7 Established computer-based cognitive assessment

A brief test battery in Psytoolkit [9] was saved for all participants to perform. The test battery included two tasks, simple response time and choice response time respectively for all participants. In the simple response time task, participants react to a single stimulus by pressing one possible input on the keyboard. The stimulus is a black ‘x’ which appears in a white checkbox. Participants are instructed to respond by pressing the space bar as soon as the the ‘x’ appears. Upon providing the correct keyboard input the task resets, displaying an empty white checkbox and the cycle repeats with unpredictable delays to stimulus presentation. The choice response task operates similarly but, requires input to one of four possible stimuli by pressing one of four corresponding keyboard inputs. Four white checkboxes are displayed at the centre of the display and the two leftmost boxes correspond to input from adjacent keys ‘z’ and ‘x’ while the two rightmost boxes use adjacent keys of the comma and full stop. Before starting either task, the user is given written instructions and several practice trials with feedback.

### 2.8 Analysis of high-speed video footage

Data was reduced from slow-motion video footage of participants performing trials of the PRT. Elapsed time in video can be calculated if the number of frames present/captured between two events and the frequency of still frame capture (frame rate) set by the camera is known. In our experiment, footage was recorded at a rate of 240 frames per second, therefore, a still frame is captured every 4.1667 milliseconds. The footage provides visible confirmation of both LEDs displaying red light (randomised delay) and the transition to providing choice stimulus (green light at one LED) to the user. The first frame where the choice stimulus becomes apparent is therefore the reference frame and the number of frames following describe the elapsed time from stimulus presentation. A criterion for defining and marking events of movement in the video footage which can be compared to measurements of the PRT was required. Examples of still frames and their movement criterion are shown in Figure 3. Four key events of movement response were chosen - two events at the location of the starting foot position (first measurement obtained by the PRT) and two events at the finishing foot position (second PRT measurement). The rationale of how key events were defined was to critically assess the recording accuracy of the PRT for use in research and practice settings. For example, the criterion of key event labelled A - Initiation (Figure 3) is the first frame which displays any sign of movement - a visible deflection in the position/shape of the laser on the user’s foot. The next event, labelled B - Lift-off, has criterion of the first frame displaying complete clearance between the user?s foot and artificial turf. Considering how measurements are obtained at the sensory circuits of the PRT, the key event A - Initiation identifies the first signs of responsive movement at the user’s feet but, not enough movement to have the sensory circuit unobscured. Conversely, the event B - Lift-off describes an event where the movement response has advanced and the sensory circuit, ±0.5cm above the turf, could be partially exposed to light or unobscured light existed while a part of the foot continues to break contact with the turf. It was therefore hypothesized that measurements obtained by the PRT would lie within the window of key event A and B indicating strong agreement between elapsed time calculated from video footage and measured by the PRT. Thereafter, the application and clinical accuracy of the PRT can be assessed by judging the magnitude of the time frame between event A and B.

**Fig 3.**
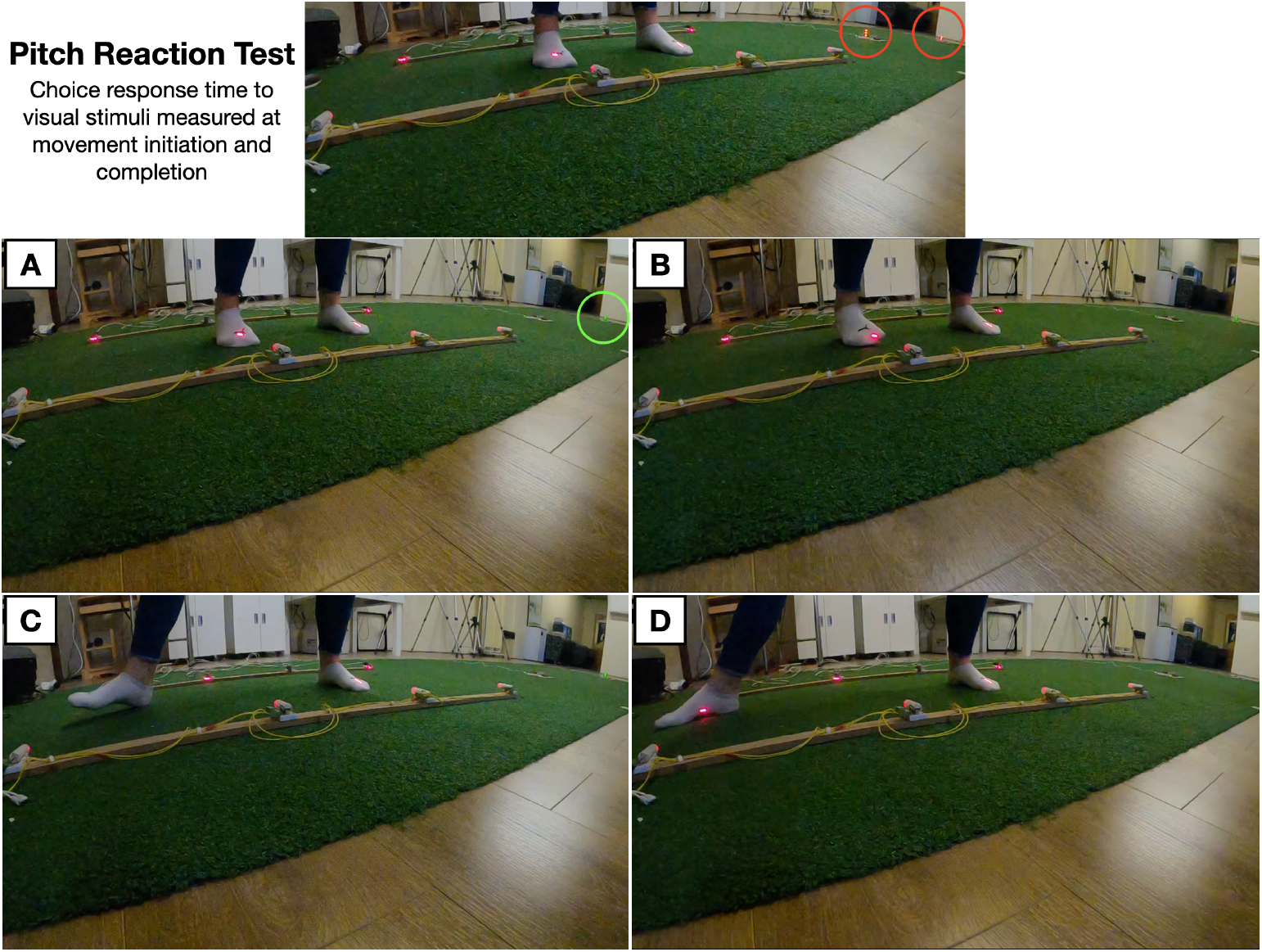
Outline of key events during one trial of the Pitch Reaction Test. Top unlabelled image depicts the sensory preloading phase where both stimuli/choices pulsate the colour red for a randomized time delay before randomly presenting a ‘go’ signal (green light) at one of the available choices - forward foot or back foot. Image A depicts Initiation of movement, the first frame with visible deflection of the laser on the athlete’s foot after being presented the go signal. Image B depicts movement Lift-off, the first frame with visible clearance between the athlete’s foot and artificial turf. Image C depicts Point of Contact, the first frame with visible contact of the athlete’s foot with the artificial turf at the position of correct choice to the stimuli presented - in this case stimuli presented at back light requires back-foot movement. Image D depicts end of movement or Finish, the first frame where the athlete’s foot makes complete contact with the turf. The go signal is deactivated once the PRT detects a correct response and the cycle repeats for subsequent trials.

### 2.9 Data Analysis

Each participants performed a total of 15 trials in the assessment. All data were tested for homoscedascity from Q-Qnormal plots and passed Shapiro-wilk test for normality. Multiple linear models assessed the strength of relationship between measures obtained from the PRT and calculated at each key event from video footage. Pearson correlation coefficients (r) were also calculated for each key event. Bland-Altman analysis is also performed to determine the mean difference and limit of agreement between the two methods at each key event. From mean values, linear models assessed the relationship between the mean response time achieved on the PRT by each participant against the mean response time achieved on the simple and choice reaction tasks of PsyToolkit.

Finally, linear models of the relationship between participant height and true leg length with mean PRT response times are performed.

## 3 Results

Anthropometric variables of 23 recruited participants are summarised in Table 1.

**Table 1.** Height and true leg length of male and female participants.

|  | Male (n = 10) |  | Female (n = 13) |  |
| --- | --- | --- | --- | --- |
| Height (cm) | 171.5 | (7.3) | 165.7 | (5.5) |
| True Leg Length (cm)<sup>1</sup> | 89.3 | (6.4) | 86.5 | (4.5) |

Data presented as mean (SD)
<sup>1</sup>True leg length measured from iliac crest to medial malleolus according to method of [8]

### 3.1 PRT comparison to high-speed camera

Elapsed time between presentation of stimulus and primary movement response measured by the PRT and high-speed camera were correlated at A - Initiation (*r* = 0.990) and B - Lift-off (*r* = 0.991). Similar, measures between stimulus presentation and final movement were also correlated at C - Point of Contact (0.975) and D - Finish (0.970). Figure 4 compares elapsed milliseconds recorded by both devices at each interval, with a line of perfect agreement (slope = 1, intercept = 0) and calculated linear models presented in Table 2.

**Table 2.** Co-efficients of agreement and correlation between the PRT and high-speed camera at each key event of movement.

| Event | $\beta$ | t-stat | Pr ($> \vert t\vert $) | R<sup>2</sup> | p- value |
| --- | --- | --- | --- | --- | --- |
| Initiation | 1.14 | 99.95 | ¡0.001 | 0.98 | ¡0.001 |
| Lift-off | 1.14 | 104.86 | ¡0.001 | 0.98 | ¡0.001 |
| Point of contact | 1.11 | 63.03 | ¡0.001 | 0.95 | ¡0.001 |
| Finish | 1.13 | 55.61 | ¡0.001 | 0.94 | ¡0.001 |

R<sup>2</sup> : coefficient of determination

**Fig 4.**
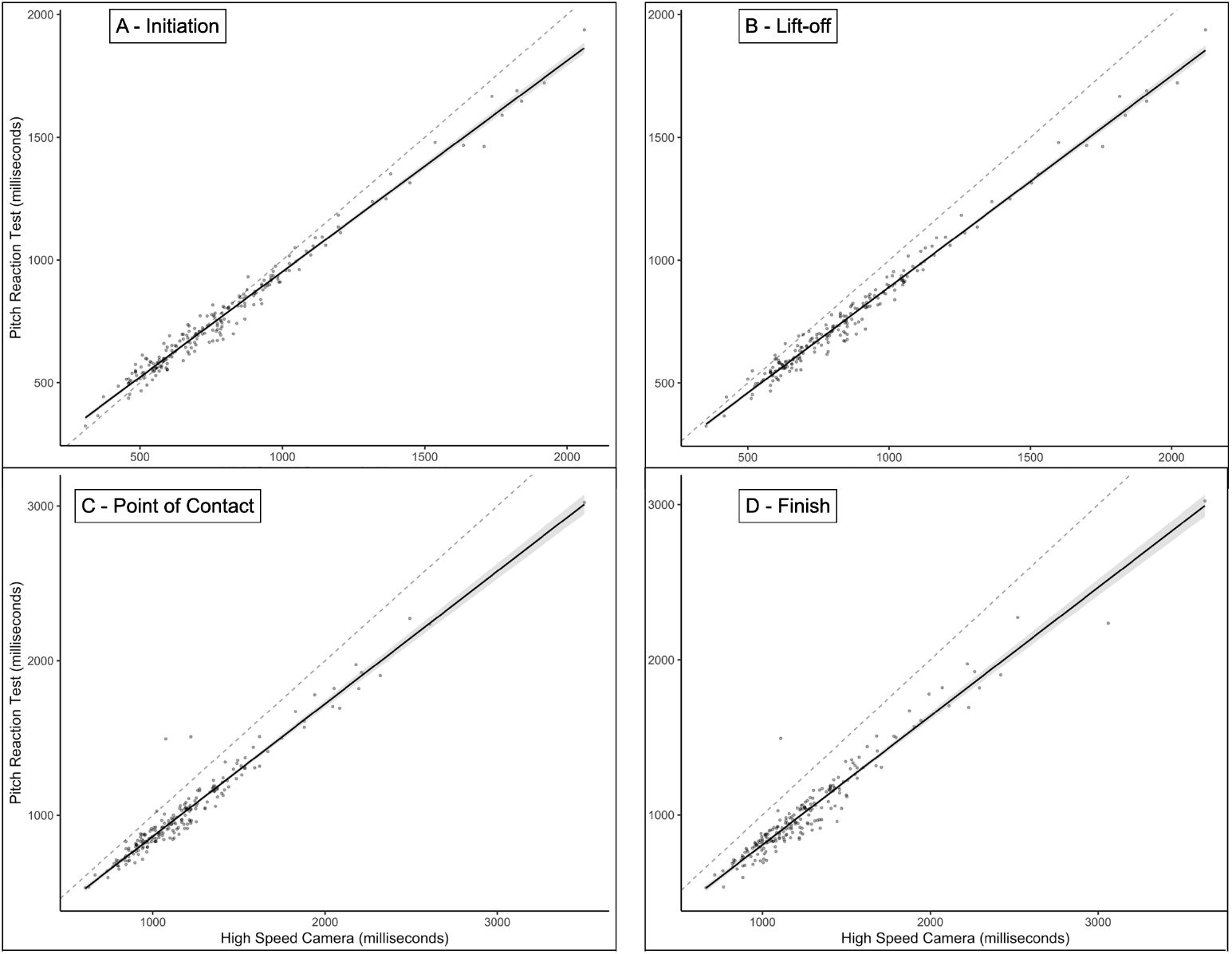
Linear models comparing measurements calculated at each key event of video footage with measures obtained from the PRT. Solid line indicates applied model and grey shaded area represents the 95% CI. Dotted line represents a line of best fit with intercept = 0 and slope =1

In the Bland-Altman analysis, the mean difference between measures increased from 17ms measured at movement A - Initiation, to 89.4ms at B - Lift-off, 162.8ms at C - Point of Contact and 236.2ms at movement D - Finish (Figure 5).

**Fig 5.**
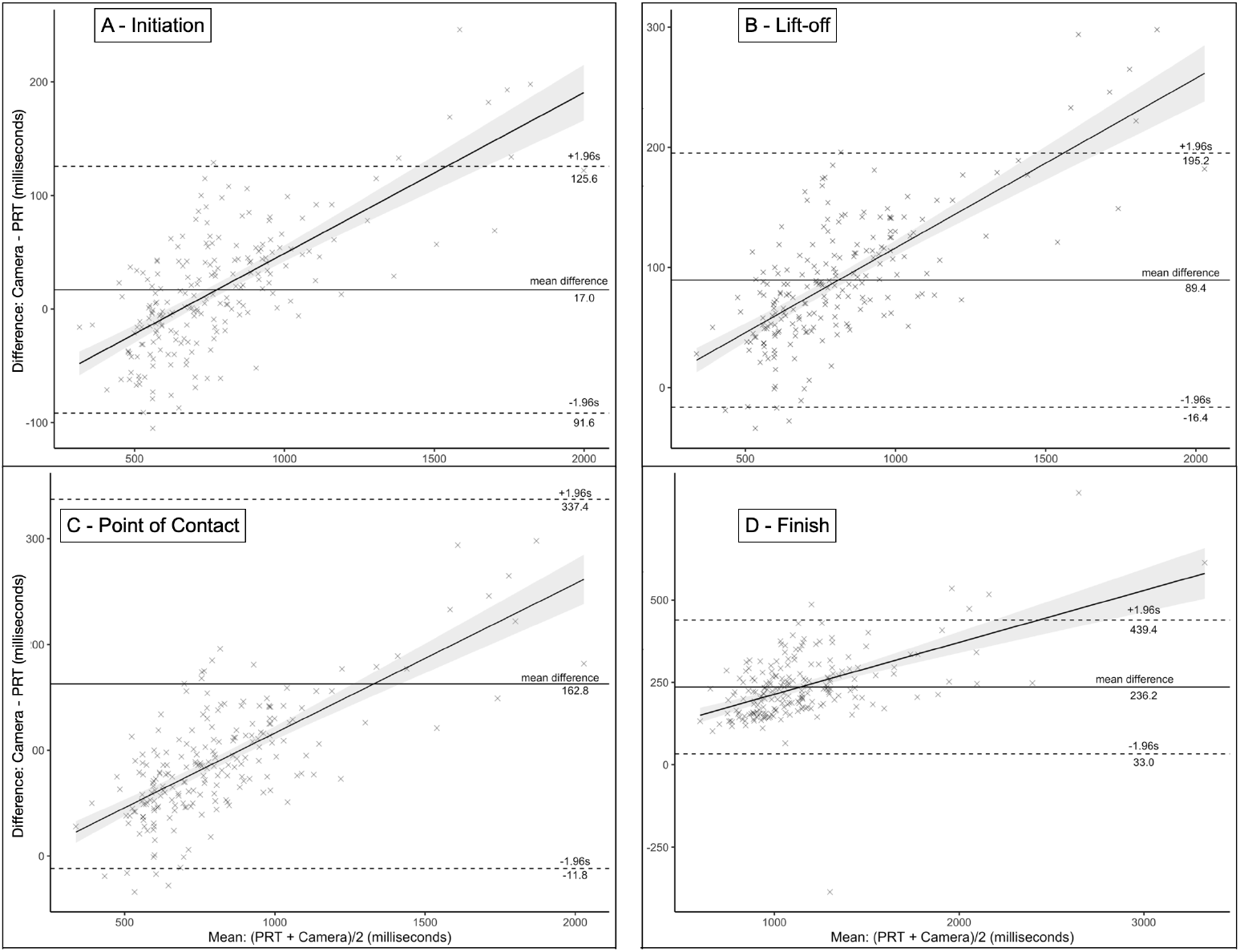
Bland-Altman analysis comparing mean difference and limits of agreement at each key event. Dotted lines represent upper and lower limits of agreement and solid horizontal line represents mean difference. Overlaid linear models with 95% CI shaded in grey are also applied showing linear relationship between mean response times and differences between methods

### 3.2 Correlation with subject anthropometry and equivalent seated task performance

Choice response times at the computer-based task showed a weak (*R*^2^ = 0.16, *F* = 4.61, *p* = 0.04) negative relationship to PRT performance (*β* = −0.21, *t* = −2.15, *p* = 0.04). One item response times were not predictive of PRT performance (*β* = −0.07, *t* = −1.44, *p* = 0.17) - Figure 6. Neither subject Height (*β* = −0.01, *t* = −0.63, *p* = 0.54) or Leg Length (*β* = 0.00, *t* = −0.56, *p* = 0.58) were significant predictors of PRT performance (Figure 7).

**Fig 6.**
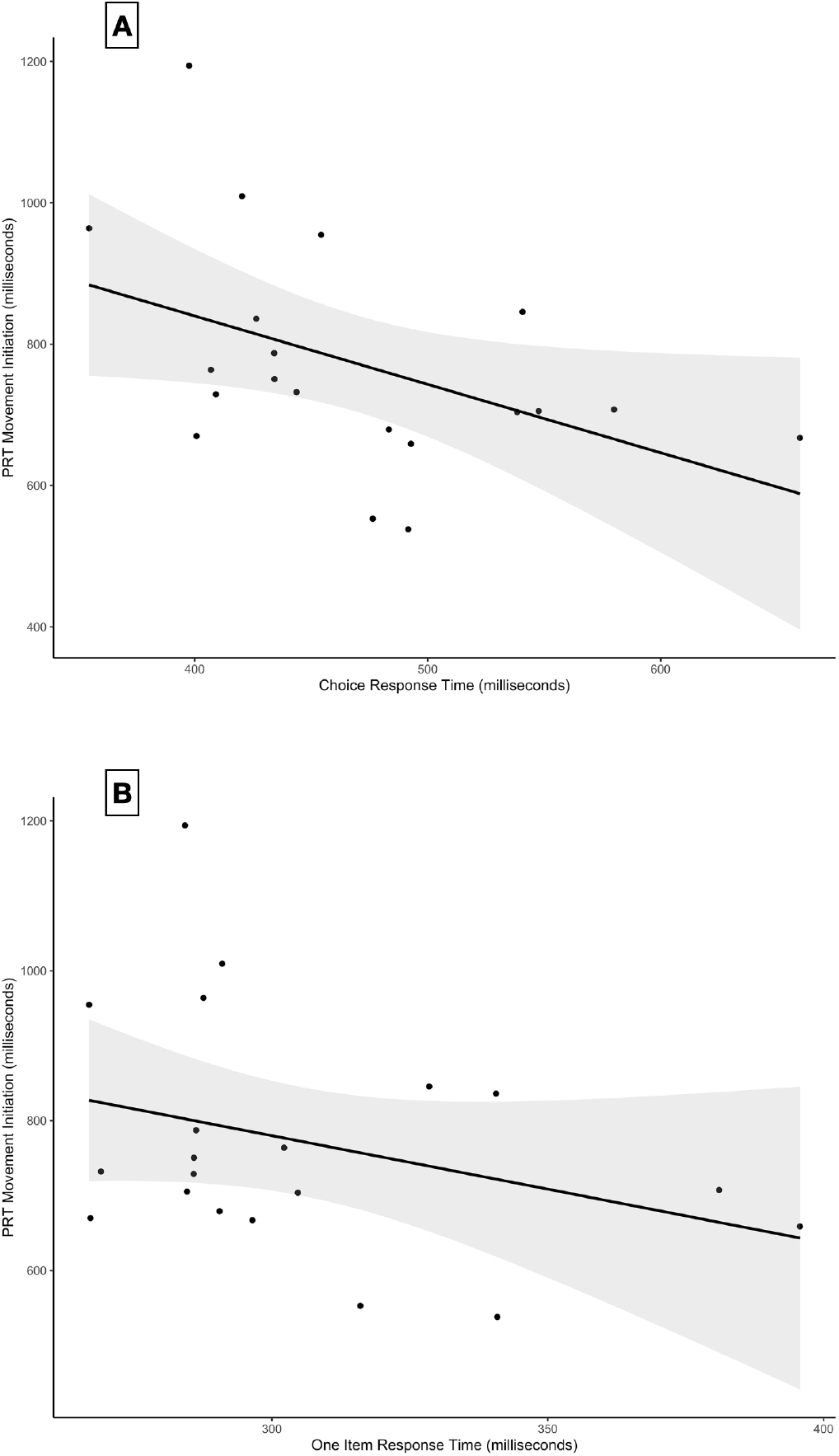
Linear models between PRT performance measured at the first sensory array and computer based analogue assessment (PsyToolkit). A - depicts relationship with choice response task consisting of four potential stimuli and four corresponding inputs. B - depicts relationship with single item task of one stimulus and one input

**Fig 7.**
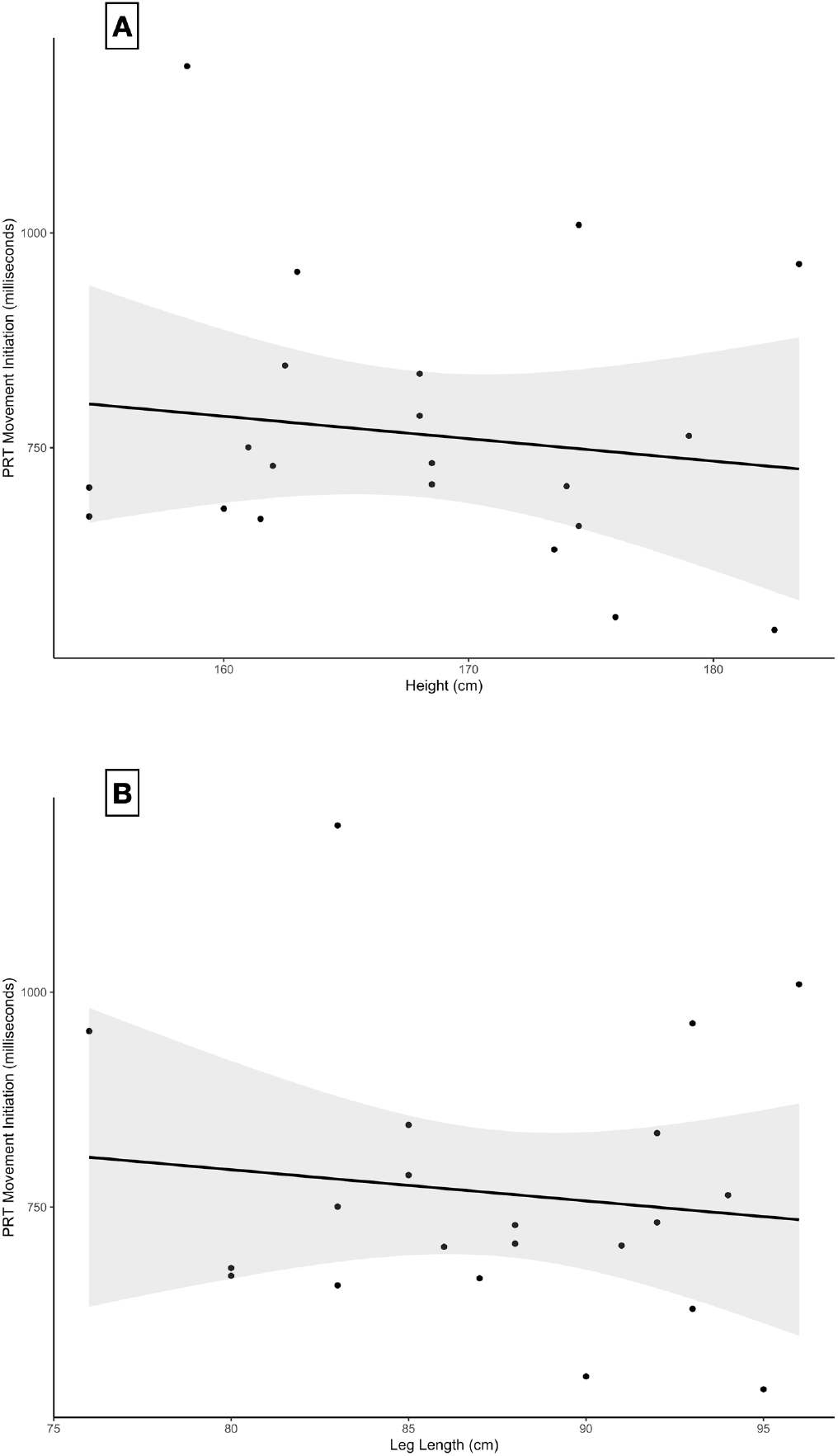
Linear models between PRT performance and subject anthropometry. A - depicts relationship with subject height. B - depicts relationship with true leg length measured according to [8]

## 4 Discussion

The developed PRT demonstrates a practical solution to measuring the speed of cognitive, psychomotor function in cricket players in the playing environment. Measurements calculated from video footage and the PRT are strongly correlated, although a deviating trend is visible. Height and leg length do not appear to be strong predictors of PRT performance. There’s a weak negative relationship between choice response times on the computer-based assessment and the PRT.

The measurements obtained by the PRT show a strong relation to those calculated from video footage. At all four key events, the PRT is correlated to within *r* = 0.98 − 0.99 of the high-speed camera (Figure 4). Linear models, however, do not show a perfect relationship of gradient and intercept or line of best fit (gradient = 1, intercept = 0). Instead, coefficients range from *β* = 1.11 − 1.14 indicating that the PRT records measurements which are smaller in magnitude i.e., faster response times by 2-19%. The trend is further clarified by Bland-Altman analysis at each key event. The mean difference between measures obtained by the PRT and high-speed camera is smallest at event A - Initiation (17ms) and thereafter increases at each subsequent event till a difference of 236ms is observed at event D - Finish (Figure 5). Further, differences between the PRT and high-speed camera increase linearly within each key event (Figure 5) as the mean response time increases. In other words, slower responses produce less agreement and greater difference between methods.

The progressive increase in disparity therefore shows that methods initially diverged, and accuracy is steadily lost, although precision/consistency in measurement is present (Figure 4). Considering the strong linear relationship between methods, if it is assumed that the high-speed camera method obtains more accurate measurements, PRT measures can be recalculated to increase by *β* = 1.1 − 1.14 times to reflect the camera. We postulate that the high-speed video method steadily loses accuracy due to variability in still frame capture leading to accumulation of error. That is, at 240fps in 1000ms a still frame is captured on average every 1000*/*240*ms*. Therefore, the first still frame to capture the green light stimuli displayed by the PRT is ±4.166ms imprecise to the true moment of appearance. Subsequent still frames continue to approximate by ±4.166ms and resultantly error accumulates as a function of multiplying over several still frames. Introducing more still frames then, as expected at the latter events of movement (C, D) or slower responses, will result in more accumulation of error and less agreement between methods.

A device like the PRT is undoubtedly more practical than measuring response times to visual stimuli from high-speed video footage. Although the true accuracy to reality cannot be fully concluded with the data and method presented in this study. The components of the PRT have a theoretical sensitivity of ±30ms which should supply sufficient resolution to defer clinical significance changes in response time performance [10]. The PRT shows potential for use in research and practice where measuring acute and longitudinal changes to response time performance within an individual are valuable. To further explore the utility the PRT, test-retest reliability should be assessed.

Given the sparsity of data surrounding the cognitive faculties of batters, a quantitative assessment of cognitive performance which is deployed on the field can aid in answering several hypotheses. Studies have shown that the effects of physical exertion on cognition quickly fade when activity is stopped [11, 12]. A device like the PRT allows parameters of cognition to be assessed concurrently with a prolonged, fatiguing batting innings. More parameters of cognition could also be considered in the test battery of the PRT. For example, although not fitting within the scope of this study, the developed python and Arduino scripts also provide a means to assess response inhibition [13]. In the task paradigm of Go/No Go tasks, stimuli can be classed into responding stimuli (single green light in this scenario) and non-responding stimuli. Non-responding stimuli somewhat mimic the appearance of responding stimuli but require executive control to inhibit the accustomed response to prepotent stimuli [13]. In our scenario, the combination of Python and Arduino code provide function for a non-responding stimulus to be delivered randomly in 20% of trials. Following the normal priming sequence and randomised delay, a mimicking stimulus (non-green light at one LED location) is delivered, and no movement response must be seen. The minimal addition to code enables two categories of tallying errors (accuracy) to be complimentary measured with response time (speed). A movement response to a green light, but to the incorrect direction is classified as a ‘legal error’ error and any movement response to the mimicking stimuli ‘inhibition/commission’ error [14]. These error classifications may supply additional insight to higher-order, executive processing performance [14, 15].

On sport performance, a similar device (Batak Lite test; [16]) requires athletes to respond to visual stimuli presented arranged in an ‘X’ pattern of the 1.1m x 1.8m grid. [16] claim that the device can be used to improve hand-eye coordination, reaction times and endurance. Veness et al., [17] induced mental fatigue in elite cricket players and the mentally fatiguing condition did not negatively affect performance on the Batak Lite test. But other cricket specific performance measures (e.g., running between the wickets) were negatively affected [17]. The PRT could be similarly used to assess mental fatigue or even provide an alternate means to induce mental fatigue compatible with batting.

## 5 Conclusion

The accuracy of the PRT appears well correlated with measures obtained from highspeed video footage. The precision of measures may require calibration to reflect measurements from video footage, however, error is predictable and consistent, which indicates acute changes in response times can be reliably quantified. Although further validatory studies of test-retest reliability are necessary. More pertinent, is the current lack of quantifiable measures of cognition which can be concurrently obtained with batting and the PRT appears a credible solution. Response time performance of novice PRT users appears inversely related to standardized, computer-based analogues which further suggests a stand-alone, cricket specific task may provide relevant and novel data to the sport. Future research should identify methods of replicating other affordances of batting behaviour to measure decision making speed from the primary motor actions of stroke play.

## Data Availability

All data produced in the present study are available upon reasonable request to the authors

## Supplementary information

Not Applicable

## Acknowledgements

Not Applicable

## Declarations

The authors have no competing interests to declare that are relevant to the content of this article.

